# Strategies for sustaining sexual and reproductive health services during recurrent flooding in Adamawa State, Nigeria: a qualitative study

**DOI:** 10.64898/2026.10.01.26364542

**Authors:** Grace W. Kimemia, Ibrahim Balami, Naoko Kozuki, Aisha Jaafar Abubakar, Jummai Umar, Maisaratu A. Bakari, Hashiya Shuaibu Muhammed, Francis Junior Ojemu

**Affiliations:** International Rescue Committee, Nairobi, Kenya; International Rescue Committee, Maiduguri, Nigeria; International Rescue Committee, New York, NY, United States; Modibbo Adama University Teaching Hospital, Yola, Nigeria; Adamawa State Government Hospital Management Board, Yola, Nigeria

**Keywords:** community-based service delivery, maternal healthcare access, climate-related emergencies, humanitarian health systems, health system resilience

## Abstract

Seasonal flooding in Adamawa State, Nigeria damages health facilities, breaks supply chains, and displaces health workers and communities, while increasing health needs. Sexual and reproductive health (SRH) services are often deprioritized, yet evidence on how they are sustained remains limited. We examined how SRH services were sustained during recurrent flooding. We conducted a qualitative descriptive study in the flood-prone Local Government Areas of Numan and Fufore. We held eight focus group discussions with women and girls and 20 key informant interviews with health providers, supply chain actors, and program implementers. Flooding closed or damaged health facilities, broke supply and cold chains, and displaced health workers. Participants linked these disruptions to unattended births and to rising hemorrhage, stillbirth, miscarriage, and genital infections. Service continuity depended less on the formal health system than on anticipatory, community-led adaptation across three phases. Before floods, stakeholders mapped risk, pre-positioned commodities, trained women in contraceptive self-injection, and dispensed medicines in advance. During floods, care shifted to mobile outreach, task-sharing, transport by canoe and boat, and community-led referral. Community health workers, traditional birth attendants, Ward Development Committees, and women’s groups reached cut-off communities. Across seasons, adaptations evolved toward cash transfers, savings groups, local reusable-pad production, and decentralized commodity hubs. Continuity relied heavily on unpaid community labor and women’s own resources. Sustaining SRH services in flood-prone settings requires institutionalized preparedness, decentralized supply, empowered communities, and protected health financing. Community-led models offer a transferable approach to health security in a changing climate.

## 1 Introduction

Climate-related shocks disrupt essential health services in low- and middle-income countries, where health systems are often resource-constrained and poorly prepared to absorb a sudden rise in need (1). Flooding damages health infrastructure, interrupts supply chains, and displaces both health workers and the populations they serve. Sexual and reproductive health (SRH) services are especially exposed, because they are often deprioritized in the acute phase of an emergency even though the need does not fall during a crisis (2). Climate shocks also expand existing gender inequalities as they restrict women’s mobility, add to their caregiving burden, and heighten the risk of gender-based violence in displacement and shelter settings (2–4). The analysis by MSI Reproductive Choices across 26 climate-affected countries estimated that some 11.5 million women had their access to contraception disrupted by climate-related displacement between 2011 and 2021, with a further 14 million at risk over the following decade unless access is protected (4).

Sub-Saharan Africa combines high exposure to climate shocks with pre-existing gaps in SRH provision and limited capacity to absorb disruption. The combination places women and girls in the region at particular risk(4,5). In Nigeria, the 2022 floods affected 34 of Nigeria’s 36 states, displaced more than 1.3 million people, and left over 600 dead (6,7). Flooding in 2024 displaced hundreds of thousands more across the northeast (8). Adamawa State, in the northeastern Nigeria, experiences recurrent seasonal flooding, making flooding a predictable annual disruption.

Despite this, the existing literature focuses on documenting the harm and disruption that flooding causes, with less attention to the strategies used to ensure continuity of SRH services across recurrent floods (9). In Nigeria, prior work has examined anticipatory cash transfers and the wider public health consequences of flooding, such as cholera and malnutrition (10). A recent qualitative study in Borno State documented how the 2024 floods disrupted maternal and child healthcare, damaging health facilities, interrupting antenatal care and immunization, and destroying medicines and cold chains (11). The study documented the scale of disruption but did not examine how services were sustained.

Evidence on continuity of SRH services comes mainly from other fragile and humanitarian settings, where community health workers (CHWs) sustain essential services during crises (12), self-care interventions such as self-injectable contraception support continuation where health facility access is disrupted (13,14), and community-led and client-level strategies (15,16). These strategies are the practical adaptations on which service continuity currently depends, and the foundation on which more deliberate and better-resourced responses could be built. This study, therefore, examined the strategies through which SRH service access and provision could be sustained during recurrent flooding in Adamawa State, Nigeria, drawing on the accounts of women and girls, health providers, supply chain actors, and program implementers.

## 2 Materials and Methods

### 2.1 Study design and setting

We adopted a qualitative descriptive approach, using focus group discussions (FGDs) and key informant interviews (KIIs) to examine how SRH services were accessed, disrupted, and sustained during flooding.

Adamawa State lies in northeastern Nigeria and experiences seasonal flooding along the Benue and Gongola rivers, with severe events recurring in recent years. We purposively selected two flood-affected local government areas (LGAs), Numan and Fufore, for their repeated exposure to flooding, and within them four communities, each served by a primary health care (PHC) facility: Imburu and Pare in Numan LGA, and Fufore and Daware in Fufore LGA. Data were collected in August 2025.

### 2.2 Study population and sampling

We purposively recruited women and girls, health providers, and supply chain actors, and used snowball sampling to reach program implementers. Women and girls of reproductive age (15 to 49 years) living in the study communities took part if they had experienced disruptions in accessing SRH services during or after flooding. They were recruited through the study PHC facilities and community mobilizers.

Health providers delivering SRH services at the study facilities, including midwives, nurses, and CHWs, were identified at each PHC through their role in service delivery. Supply chain actors were those responsible for the SRH commodity supply chain, selected to trace the chain from the state central medical store through the LGA to facility level. Program designers and implementers of flood-related adaptive strategies were drawn from governmental and non-governmental organizations operating in Adamawa State, and in some cases more widely across north-eastern Nigeria. They were identified from an initial list compiled by health and economic recovery and development technical specialists in the International Rescue Committee, Nigeria office and expanded through snowball sampling.

We conducted eight FGDs with 63 women and girls in total, stratified by age and community to capture SRH needs across the four affected communities. In each of the four communities, we held one FGD with adolescent girls and young women aged 15 to 24 years and one with women aged 25 to 49 years, each group comprising six to ten participants. We conducted 20 KIIs, comprising eight health providers (two per facility), seven supply chain actors at state, LGA, and facility levels, and five program implementers. Sampling continued until the data reached saturation, the point at which further interviews and discussions yielded little new information relevant to the research questions.

### 2.3 Data collection

Data were collected by trained female field researchers experienced in qualitative methods. The field team completed a three-day training covering the study objectives, research ethics, the informed consent process, and the data collection tools. The training was followed by two days of piloting the tools. We used semi-structured guides for the FGDs and KIIs, with separate guides tailored to each participant group and developed around the research questions across the flood cycle. Each session was conducted in the local language of Hausa or English, according to the participant’s preference, at a private and mutually agreed location such as a health facility or a community venue.

FGDs and KIIs were audio recorded with consent, transcribed in Hausa or in English, and the former translated into English. Field researchers also kept notes during data collection. Recordings and transcripts were stored in password-protected folders accessible only to the research team.

### 2.4 Data analysis

We organized the data using a data stripping approach, a structured matrix method in which each participant forms a row and the columns hold their characteristics and the interview topics (17). Transcript responses were entered into the relevant cells, and short analytical notes on the general trend and on the variation observed were recorded at the foot of each column. Where participants raised issues that fell outside the interview questions, we added columns to the sheet to capture these emergent themes. We then analyzed the charted matrix thematically, developing and refining themes across the three flood phases. The research team members (FJO, IB, AJA, JU) carried out this process, and the lead authors reviewed the resulting summaries against the underlying transcripts to confirm that the interpretation was consistent with the data and to resolve differences through discussion.

### 2.5 Ethical considerations

We obtained ethical approval from the International Rescue Committee Institutional Review Board (H.1.00.087) and the Modibbo Adama university Teaching Hospital, Yola Health Research Ethics Committee (HREC) (MAUTHY/SUB/S.128/404). All participants provided written informed consent before data collection. Participants were assured that their decision to take part would not affect their access to or quality of care or, for health providers, their employment. Both review boards waived parental consent for married adolescents aged 15 to 17 years. This reflected the sensitivity of the topics and the recognition of married adolescents in this setting as emancipated.

## 3 Results

We present the findings in three phases relative to flood onset: before flooding, during active flooding, and across successive seasons.

### 3.1 Strategies before flooding

Before or at the onset of flooding, stakeholders described anticipatory strategies centered on identifying who and where would be most affected, protecting and positioning commodities, and equipping women to manage their own SRH needs through the disruption. At the LGA level, supply chain actors described how the executive secretary; who has oversight functions for all PHCs in the LGA, under the Adamawa state Primary Healthcare Agency, mapped settlements at greatest risk and redirected stock and staff accordingly. As one supply chain reported that *“our Executive Secretary directed the LGA staff to shortlist settlements at greater risk… he assigns the little stock we have in the LGA to be redirected to those locations*” (KII, supply chain actor, SC04).

Health facilities, on the other hand, extended this mapping to individual women, listing pregnant women and women of reproductive age in flood-prone areas. This mapping allowed anticipatory distribution to be targeted before access was lost. As the rainy season approached, health facilities reviewed their records to identify the conditions that recurred during floods. They submitted early requisitions to the LGA for the relevant medicines and kept a rotating buffer stock of fast-moving commodities. They also adjusted staff duty rosters for the anticipated disruption.

Before the rainy season, stakeholders convened joint planning meetings to agree on stock levels and arrangements for service continuity. These meetings brought together PHC staff, Ward Development Committees (WDCs), LGA authorities, and implementing partners, as one supply chain actor described:

…*before the floods arrive, we meet at the Ward Health Quarters… we already plan to make sure enough medicines are in stock. All primary healthcare centers have adopted proactive strategies… such as pre-positioning essential supplies in elevated or secured storage locations.* (KII, supply chain actor, SC06)

Some health facilities trained emergency response teams in advance and visited communities before the floods, as one supply chain actor narrated:

*We visited villages before July, August, and September… and told them when the floods come there will be no road to the facility… If possible, we left him with some drugs like paracetamol or antimalarial, and explained how to give them.* (KII, supply chain actor, SC06).

Pre-positioning was among the most common measures. At the state level, supply chain actors released larger than usual quantities of medicines to health facilities likely to become inaccessible. In some instances, health facilities used solar-powered refrigerators to protect oxytocin and vaccines when power failed. One supply chain actor described, “w*e need to ensure that there is adequate stock… especially if there is flood prediction. We try to issue more than what we actually issue during normal cycles to such location.”* (KII, supply chain actor, SC07). At the LGA level, the health facilities were prompted to secure their own stock ahead of the flooding as quoted, “*when the flood is about to come, I ask them to make arrangements for where they will keep their drugs so that the drugs will not be affected by the flood*” (KII, supply chain actor, SC03).

Where movement was expected to become impossible, some health facilities pre-positioned drugs a step further out, into the communities likely to be cut off. In some cases, health providers left a small stock of basic medicines with CHWs in communities likely to be cut off. One of the supply chain actors mentioned that:

*When we expect that movement might become difficult, we sometimes pre-position drugs in nearby communities that are usually cut off by the flood… even if roads become impassable, community health workers in those areas can still provide basic care and report back to us (health providers)* (KII, supply chain actor, SC02).

The composition of pre-positioned stocks reflected awareness of flood-specific SRH risks, as one supply chain actor narrated, “*during the flood period, we make sure to stock specific sexual and reproductive health commodities… Items like HIV test kits and other sexually transmitted infection diagnostic kits are always prioritized because, during such times, the risk of sexual assault and unprotected sexual contact tends to increase. We also ensure the availability of PEP kits for managing potential HIV exposure and emergency contraceptives for cases of unintended pregnancies*.” (KII, supply chain actor, SC02)

These facility-level efforts were reinforced by planning at higher levels. At the state level, SRH commodities were written into the annual operational plan with reserved budget. This was an effort to protect supplies through routine planning. Health facilities also documented consumption systematically to improve forecasting from one season to the next, *“we started taking proper records of the number of victims we supported and the types and quantities of drugs they used… so we planned for more paracetamol, antimalarials, and ORS*” (KII, supply chain actor, SC05). This documentation showed needs for which SRH commodities surged during floods, which helped health facilities anticipate need more accurately.

Health providers also prepared women to manage their own needs through the disruption. For family planning, women were often given a full year’s supply of injectable contraceptives alongside training to self-administer at home, as one health provider narrated, *“we give them for a year… they will use it after every three months… we will guide you as we watch, then you will do it to yourself. Then we give you the remaining three and you take them home with you.”* (KII, health provider, HP07)

For pregnant women, health providers dispensed antenatal care (ANC) medicines such as iron folic acid and malaria prophylaxis ahead of the flood season so that monthly supplies could cover the period when access would be lost: “*for pregnant mothers that do come for antenatal, if they normally receive drugs on monthly basis, they will be given for two months so that whenever there is a lack of access road to meet the service providers due to the flood the drugs can cover*” (KII, health provider, HP07). Traditional birth attendants (TBAs) were also provisioned with medicines in advance so they could support pregnant women in cut-off areas. Health providers also trained women in home-based sexually transmitted infection (STI) testing where kits were available.

Women also prepared before flooding. Knowing the flood calendar, they purchased and stored the medicines they expected to need, including contraceptive supplies, before access was lost, as one participant described that, “*I do collect and keep my child’s medications, the ones I would use such as pills that women do use, to store because even when the flood came, I wouldn’t have clinical problems to visit.”* (FGD02, women group)

Others visited health facilities to purchase medicines ahead of time, and some described keeping a personal emergency SRH kit at home: “*having personal emergency SRH kits stored in the house and staying informed about where to access help even during disasters*” (FGD07, adolescent girls and young women).

Anticipating menstrual needs was also part of this preparation. Program implementers pre-positioned dignity kits in accessible locations before the water rose. They also advised women and girls to buy sanitary supplies ahead of the season while markets remained reachable, since these would be hard to obtain once access was lost. Women who had received self-care training reported drawing on it during floods. Those without such training, or without the means to stock up in advance, they had fewer alternatives when health facilities became inaccessible.

### 3.2 Strategies during active flooding

When flooding disrupted access to health facilities and displaced populations, stakeholders described operational adaptations to maintain SRH service delivery. These involved taking services to displaced women, keeping commodities and staff moving, and drawing on community actors at the last mile. However, women’s accounts also revealed the limits of these adaptations.

One of the most significant operational shifts was the move from static, facility-based care to mobile and community-based delivery. When health facilities became inaccessible or populations were displaced with shelters and schools, health workers followed displaced populations and set up temporary clinics at accessible locations, as one health provider described:

*During the flood disaster, we made several adjustments for women and girls. We brought services closer to affected communities through outreach visits and mobile health activities… We also formed and trained community health support groups made up of two to three individuals from each area, equipping them with basic health knowledge. These volunteers helped identify pregnant women and girls in need of care… we arranged for boat transportation to refer patients to specialist hospitals.* (KII, health provider, HP06)

Community health support groups were widely used. They connected health facilities with their communities, offering basic counseling and alerting staff to emergencies. Since repeat trips into cut-off communities were rarely feasible, outreach was deliberately integrated so that women received the full range of services in a single visit: “*If we go out, we go out integrated, everything is available, we provide antenatal and postnatal care, immunizations, HPV testing, and other services we do them all*” (KII, health provider, HP03).

Where flooding was severe, Executive Secretaries could direct PHCs to relocate, as one supply chain actor described: “*If PHCs are affected… the staff work as outreach clinic and follow those people displaced by flood to their new settlement”* (KII, supply chain actor, SC04). To maintain privacy for intimate care when shelters were crowded, health facilities established women-only consultations as one supply chain actor explained: “*He (the community leader) gives us a space where women are attended to separately to ensure privacy. Female health workers provide health care services*” (KII, supply chain actor, SC04). For deliveries, health providers discouraged unsafe home births where referral was possible and arranged transport to the nearest functional PHC.

Stakeholders reported that sustaining the workforce required flexibility. Health facilities reorganized staff roles, designating focal person and reassigning duties when individuals could not safely cross floodwaters. One of the key informants described, “*we divided ourselves, and everyone was assigned a specific responsibility… the ANC in-charge handles ANC, labor, and delivery. If she has any need, she tells me and I take the request upward.”* (KII, in charge, SC01)

Staff safety and protective equipment shaped how much coverage was possible. Health providers with rain boots and life jackets maintained broader coverage than those without, although many health facilities lacked such equipment. Health providers providing services through outreach noted that it was dangerous, saying that boats “*sometimes capsize, leading to loss of life*.” Workforce shortages compounded these constraints: “*Our workforce is too small to sustain proper 24-hour service coverage. I am the only midwife here… I work both day and night shifts without adequate* rest” (KII, health provider, HP01).

Stakeholders reported that moving commodities to the last mile required alternative transport. When roads became impassable, supply chain teams improvised transport across the terrain as described “*The rapid response team take them to the facility themselves… where water is deep we use boats, and where the water is shallow, the WDC carry the drugs on their head to the facility*” (KII, supply chain actor, SC04). Stocks kept flowing through redistribution. The State Central Medical Store tracked health facility stock status and redistributed accordingly: “*We receive regular updates… to know whether a particular facility is about to experience stock out or not, then we can redistribute from another facility that has spare quantities*” (KII, supply chain actor, SC07).

Redistribution also happened directly between neighboring health facilities, with facility in-charges calling one another to move surplus stock and documenting each transfer on inventory control cards. When normal communication channels failed, they coordinated through community radio, and written messages carried between communities by health workers. One supply chain actor described the direct transfers between facilities as quoted below:

*If another facility in the town has a large supply of a particular drug… it is my duty to call that facility manager. They will send me some of their medicine, write it down on their records, and I will also sign for receiving it… I may have medicines that they lack… So, I will also share with them.* (KII, supply chain actor, SC01)

Financing adaptations helped health facilities procure what the supply chain could not deliver. The Drug Revolving Fund (DRF), a locally managed and self-sustaining mechanism, gave facility managers immediate procurement flexibility when central supply was delayed, as one supply chain actor described:

*When I reach the health facility and find out that they don’t have drugs, I direct them to buy the drugs through their drug revolving fund (DRF) scheme… they write the drugs for my review and approval. I will reforward the list of drugs to the executive secretary of the LGA for approval.* (KII, supply chain actor, SC04)

The DRF was faster than state requisition and allowed health facilities to maintain contraceptive, delivery, and STI commodity availability. Health facilities also priced DRF medicines to draw women into formal care rather than to informal vendors, as one supply chain actor explained: “*we sell them here at a slightly cheaper rate than local chemists… if paracetamol costs 100 Naira in the chemist, we sell it for 70 or 80 Naira ($0.05-0.06) here, so people prefer coming to the facility instead of going to the chemist”* (KII, supply chain actor, SC06).

However, the DRF’s viability depended on cost-recovery through drug sale. This would collapsed when flooding destroyed livelihoods and health providers distributed free medicines without reimbursement: “*Back when we still had support… we separated a portion of medicines and gave them to the poor of poorest free of charge. But now… honestly, there is nothing at all unless the government or some organizations come in*” (KII, supply chain actor, SC06). To reach the most vulnerable women, health teams nonetheless provided SRH drugs free of charge during floods. This was supported by emergency replenishment from the state pharmacist, routed through PHCs to affected communities. Implementing partners filled remaining gaps with emergency commodities and logistics support. In border areas, cross-border coordination with Cameroon aligned commodity pre-positioning and referral pathways.

Health providers across the four communities reported a marked increase in reproductive tract and sexually transmitted infections, which women attributed to prolonged contact with floodwater. Health provider reported that in some weeks, “*in 10 women 8 will be complaining of STI and they claim it was the flood water that enter their body and cause the STI*” (KII, health provider, HP05). The drivers were both environmental and social. Displacement into crowded shelters removed the privacy, clean water, and sanitation that hygiene requires. Confinement and the collapse of livelihoods, meanwhile, increased unprotected and transactional sex.

Managing menstruation became difficult at the same moment. Women without supplies reused rags. One program implementer described how a pad would be recycled, *“one person use and wash and give it to the other,”* a practice the implementer linked to infection. Adolescent girls described genital infection as rampant, and as something that *“brings about miscarriages, menstrual complications.”*

In response, health providers treated infections syndromically where laboratory confirmation was not possible. They kept a stock of the medicines women most often needed, as one noted: *“there is a medicine for V. (vaginal) itching, because most of them come with infections”* (KII, health provider, HP08). Treating a woman while her partner went untreated returned her to the clinic with the same complaint. Health providers who could still reach communities therefore paired treatment with couple counselling, advising women who tested positive to bring their husbands for treatment. As one explained, *“because if only the woman is treated and her husband, especially one with multiple partners, is not, the infection easily reoccurs”* (KII, health provider, HP01). This couple-level and outreach work was among the first to be interrupted when flooding cut the roads, which health providers linked to a rise in untreated infections during the peak flooding weeks.

Some health providers maintained remote contact with clients, using phone numbers collected at outreach to track defaulters and to advise women who could not reach a health facility. Telephone contact was sometimes the only link to a woman in crisis, although it had limits. One health provider recounted, “*… a woman called me and said… she saw blood and she is pregnant… I asked her if there is a traditional birth attendant in her village… Whatever it is you should call me on the phone, but she didn’t call me till morning, then I searched for her contact and called her… she said she has given birth.* (KII, health provider, HP03).

Health facilities also used registers and contact lists to follow up on missed family planning and ANC appointments. These communication adaptations were not widely practiced and were constrained by network outages and the cost of airtime. However, health providers described phone contact as a helpful tool during floods.

Communities extended SRH services when formal systems failed. WDCs evolved from passive representatives into active partners, carrying medicines across flooded terrain, constructing temporary bridges, and relaying health needs to facilities: “*Sometimes we collaborate with members of the community, and they help us get the medicine from the riverbank… they know it’s for their own benefit*” (KII, supply chain actor, SC01). They also provided access routes and shelter space and advocated to local government over staffing shortages, as one health provider recounted: “*The community leaders then took it upon themselves to report the issue to the Local Government Authority, advocating for the posting of additional health workers to our facility*” (KII, health provider, HP01).

In some communities, residents organized emergency transport themselves, designating standby vehicles and drivers for women in labor: *“If someone went into labor, they would call a number, and the designated driver would come pick them up and take them to the hospital… at least one man had to remain at home for emergencies”* (KII, health provider, HP03). To keep volunteer drivers on call, some communities gave them priority access to fuel. They also issued a signed slip recording the driver’s name and the woman he had transported, which the driver could later use to claim reimbursement. Where a single vehicle could not complete the journey, transport was relayed: a tricycle carried the woman as far as the water allowed, and a canoe operator took over. Where no vehicle could pass, youths carried the woman on a stretcher. In one community, a boat was kept for hire so families could reach the health facility in an emergency.

Community structures also removed financial and diagnostic obstacles at the point of need. A WDC member could stand as guarantor for a woman who could not pay the small drug-fund charge, offering to cover it for later repayment. Mama-to-Mama support groups (trained community health volunteers) took samples in the community and returned by canoe with the results and medicines. This allowed the community to access services without a visit to the health facility.

TBAs and CHWs acted as the main extension arms of PHCs when women could not reach health facilities. TBAs held basic ANC medicines, assisted in home deliveries, and linked complicated cases to PHC staff. Health providers trained both women and TBAs in clean and safe delivery, including avoiding old razors on the cord to prevent infection. They also equipped them to manage emergencies until a skilled provider could be reached, as one health provider described:

*During floods, childbirth can be particularly challenging, so we focus on preparing women and traditional birth attendants to handle emergencies safely… we provide hand gloves for emergency deliveries and supply essential drugs to manage excessive bleeding… we train TBAs on how to handle such situations safely until a professional health worker arrives.* (KII, health provider, HP02) CHWs, including Mama-to-Mama support groups, visited households to deliver medicines, counsel on ANC and family planning, and track women due for postnatal care: “*The Mama-to-Mama group visits women in their homes… checking on pregnant women, those attending ANC or with miscarriage*” (KII, supply chain actor, SC06). Women-led groups and safe spaces functioned as parallel networks. They distributed reusable pads and dignity kits, maintained discreet stocks of emergency contraceptives, and connected survivors of sexual violence to services. When complications exceeded what TBAs and CHWs could manage, women were referred onward, often by canoe, from the PHC to the general hospital in Yola town. This chain depended on water transport being available at the moment it was needed.

When health facilities and community actors could not be reached, women turned to informal providers and to their own resources. Patent and Proprietary Medicine Vendors (PPMVs) and chemists were valued for their proximity and constant stock: “*I rather utilize the medicine store nearby, than put myself in danger”* (FGD08, women group). Some vendors lowered prices during the emergency: “*What we did was to make drugs a bit cheaper so that people could afford them… most of the workload shifted to the private facilities, because many of the government workers became less active during the flood”* (KII, health provider, HP06). Women who could not reach any health provider relied on traditional medicine and improvised treatments with limited success: “*For FP I was advice to take salt after intercourse, I take salt but later find out I am pregnant even when am not ready*” (FGD07, adolescent girls and young women,).

Beyond stocking supplies, women drew on a wide range of self-management when health facilities were unreachable. For contraception, several described calendar-based abstinence combined with negotiated withdrawal. One explained that a couple would count the days and, where they could not abstain, the husband would withdraw at ejaculation. A few used herbal contraceptives. Some women relied on breastfeeding to space births, and one participant described pretending to be sick to avoid intercourse when family planning was out of reach. Some women reported they bought condoms ahead of the season and used them while supplies lasted, one noting that “*when I was breastfeeding, I would go to get a condom so that it would prevent me from getting pregnant since I am still weaning*” (FGD06, women group), and an adolescent that “*My husband use to buy condoms*” (FGD01, adolescent girls and young women).

A herbal contraceptive whose dose set the years of protection was also described: “*You will swallow four, because I didn’t see it. But she uses it and confirm, it’s what she uses. She gives birth at the time she wants*.” (FGD06, women group). For genital infection, women turned to home remedies, most commonly herbal decoctions and sitz baths: “*…we mix turmeric, lime and cloves. When you boil and drink it, it will help by flushing the STI off your system.” (*FGD03, adolescent girls and young women). Some boiled jujube (neem), while others used henna, or sat in hot water with salt.

For menstrual hygiene, women soaked underwear in ash left overnight or in disinfectant and adjusted drying practices to reduce infection. Women also reused the self-care advice given at antenatal clinics, one describing how she had become a “*mini doctor*” at home. Where labor came, some arranged for a trusted health worker to be brought to the home by canoe. Women also proposed collective solutions of their own. These included a contributory fund through which they pooled small sums to buy infection drugs, women’s peer and gender groups for health awareness, and a named women’s support group. They further argued that women should be recognized as household planners. One participant observed that a woman would prepare her household well for the flood if the men would allow it.

Women’s own analysis located much of the infection burden in the sanitation failures that flooding produced. Above all, they pointed to the pit latrine as the pathway through which infection reached them. One explained that *“if the flood comes, it fills up the toilet and it gushes into the house… where you squat to pee you can get infection from there”* (FGD03, adolescent girls and young women). Others described contaminated water splashing during use: *“the pit toilet oozes heat and it touches one’s private part”* (FGD06, adult women). A single shared toilet could spread infection through a whole household, since *“it’s one toilet, you see? One person gets and all the household members will contact it too”* (FGD04, adult women).

In response, women adopted household hygiene practices that they shared with one another. Some abandoned the flooded latrine altogether: “*I don’t use the pit latrine again. I go back to using poo-poo bowl (a portable toilet receptacle), I wash it with hot wate*r” (FGD03, adolescent girls and young women). Others washed with salted hot water after using the toilet, believing that “*the hot water and salt will kill any kind of infection*” (FGD06, women group). They dried underwear in direct sunlight, since “*the sun removes infection*” (FGD06, women group). When boreholes were submerged, they treated drinking water, one recalling how “*you fetch the running water and put alum to purify to drink*” (FGD02, women group).

When women went into labor at home with no access to skilled delivery, some delivered with only a neighbor’s help, and others delivered entirely alone: “*I started shouting for help in my labour pains, nobody came… I gave birth at home without anybody*” (FGD03, adolescent girls and young women). One participant described a hazardous practice used in her community when no skilled help could be reached, recounting it as another woman’s experience:

*When you’re in labour at home and there is no access road… When you’re unable to push, they would bring dry chilli pepper and pour it in a open container carrying burning charcoal, the smoke generated from it will fill the room and the woman will start coughing heavily, almost non-stop, each cough is expected to help her push the baby out. The sustained cough and pain would cause the baby to come out forcefully*. (FGD08, women group)

### 3.3 Strategies across successive flood seasons

Beyond preparing for and responding to individual flood events, stakeholders described longer-term changes that emerged across seasons. These included new program approaches, community and anticipatory financing, and proposals for structural reform of the supply chain. Several implementing organizations have built gender-responsive design into their programming. This began with gender-disaggregated assessments of how flooding affected women and girls, as one program implementer described

*We place women and girls at the center of our program design and implementation. From the assessment phase, we collect gender-disaggregated data to understand their unique SRH challenges, especially during flooding when access to services is limited. Based on these insights, we design interventions that prioritize menstrual hygiene management, maternal health, and access to contraceptives*. (KII, program implementer, PI04)

Menstrual health received particular attention across seasons, addressed by different organizations in complementary ways. One organization distributed reusable sanitary pads, reaching over 350 adolescent girls alongside menstrual hygiene education in schools. Another moved from distribution toward durable local capacity, training adolescent girls to produce reusable pads themselves so that supply would not depend on access during floods: “*we were able to teach them how to produce a reusable menstrual material to produce locally within their community. We gave them a sewing machine; we gave them training and also, we trained them on how to prepare soap for themselves*” (KII, program implementer, PI05). Program implementers also described directing female-targeted interventions through women leaders rather than male gatekeepers, after women reported that relief items intended for them were sometimes diverted.

Financing strategies also emerged. Within communities, Village Savings and Loan Associations (VSLAs) functioned as a savings-based preparedness mechanism. Through weekly contributions of as little as 100 Naira (about US$0.07), VSLAs built reserves that women drew on when floods disrupted livelihoods, paying for transport, medicines, and safe delivery. However, when entire communities lost livelihoods at once, members could not maintain contributions and loan demand exceeded available funds. Some program implementers complemented these community savings with anticipatory cash transfers ahead of the flood season. The cash was deliberately left unrestricted, so that women could purchase the SRH-related items they deemed necessary, as one program implementer described:

*The resources we are giving to them as sanitary cash is not a restricted cash. We only label that you should buy everything that you need that will support you during flooding… for ladies that may require all these menstrual pads, they should buy it.* (KII, program implementer, PI05). The same transfers were paired with messaging that encouraged pregnant women to travel early to secure antenatal care and to buy clean water, linking the cash to specific SRH and maternal health goals before flooding cut off access.

In one community, a youth-led environmental initiative reduced flooding itself, and with it the access barrier that ordinarily disrupted SRH services. It did so by clearing the drainage channels whose blockage caused local flooding, as one program implementer narrated:

*For the first time, I can’t forget the (name of a community) in 2022, because of our intervention and the strategies that we had on ground, they recorded zero flooding for the first time in the history of Adamawa State… They recorded zero flooding because plastic waste became a goldmine.* (KII, program implementer, PI02)

At the supply chain level, actors articulated proposals for longer-term reform. The most notable was the decentralization of storage through zonal commodity hubs to reduce transport delays for flood-prone LGAs reliant on the central store in Yola, as one supply chain actor described:

*We are trying to see if we can get zonal hubs… at the three senatorial zones in Adamawa State… so that commodities move to a closer storage area and from there the drugs are distributed to facilities within the zones.* (KII, supply chain actor, SC07). At the time of data collection, this remained a proposal rather than implemented policy. Supply chain actors also emphasized the need for a standard operating procedure for disaster and flood logistics and for sustained funding for emergency deliveries.

Structural and system-level adaptations accompanied these proposals. Revitalized health facilities were built on raised foundations, with secured, elevated stores so that floodwater could not reach commodities. Supply chain actors also described an emerging model of direct last-mile delivery through third-party logistics, using motorcycles and cooler boxes to reach cut-off health facilities. They argued that partner supplies should be routed through the Executive Secretary’s office rather than delivered directly to health facilities. This would let a single distribution plan cover every ward and avoid parallel systems. They also proposed that anticipatory forecasting add a risk-target percentage to the normal calculation before the flood season. To protect the DRF from the perception that all medicines were free, one supply chain actor recommended sitting with community leaders to explain that some drugs are sold to sustain the fund while others are given free.

Health providers sustained services by keeping at least one health worker on duty at all times, so that emergencies could still be met. They also managed the acceptability barriers that kept women away. They offered female staff for delivery and sensitive care and let a woman wait for a different health provider or a later shift when she did not wish to disclose to the one on duty. Implementers engaged men directly to complement facility and community effort. Using equal participation quotas and advocacy so that women could take part, they reported that this had reduced conflict in households.

## 4 Discussion

Sustaining SRH services during recurrent flooding depended less on the formal health system acting alone than on a distributed set of adaptations carried jointly by health providers, supply chain actors, community structures, and women themselves. These adaptations spanned anticipatory preparation before flooding, operational improvisation during active flooding, and slower changes across successive seasons. These adaptations map onto the health-system resilience literature (1). Most were absorptive coping, through buffer stocks and improvised transport, and adaptive reorganization of how and where care was delivered. The transformative changes that would reduce the disruption itself, such as structural revitalization, zonal hubs, and sustained financing, appeared only in early or proposed form.

Much of the service continuity was borrowed, resting on the unpaid labor of communities and on women’s own knowledge and bodies rather than on resourced system capacity (12). Existing evidence on SRH in disasters and epidemics has concentrated on documenting disruption and the resumption of services afterward, rather than the strategies that keep services running through repeated shocks (9). Where adaptation has been examined, it has largely been in the context of the COVID-19 pandemic and centered on digital and facility-level responses (18).

### 4.1 Distributed continuity: community actors, self-care, and the supply chain

Community actors and women’s own capacity for self-care did much of the work of continuity. CHWs, TBAs, WDCs, and women-led groups extended primary care into communities that health facilities could not reach. This aligns with a scoping review of CHWs in humanitarian settings, which found that these cadres sustain essential services during crises but are repeatedly undermined by commodity stockouts, referral failures, and short-term funding (12). These cadres cannot, therefore, be treated as a cost-free surge capacity. Sustaining the continuity they provide means resourcing them as a standing part of the system, rather than mobilizing them unpaid each flood season. The adaptations that reached women most effectively were those that worked with prevailing gender norms. These included women-only consultation spaces, female providers for sensitive care, and routing supplies through women leaders (19).

At the client level, women received extended supplies of injectable contraceptives with training to self-administer, kept personal emergency kits, and stocked medicines before access was lost. These practices align with the World Health Organization recommendation that self-administered injectable contraception be offered as an additional delivery approach (14). A scoping review of self-care in humanitarian and fragile settings likewise found self-injection acceptable and supportive of continuation where facility access is disrupted (13). In contrast to accounts that frame self-care as an emergency fallback, we found it provisioned in advance as a planned, calendar-driven strategy for which health providers actively prepared. However, women without prior training or the means to stock up were excluded from the very strategy the system had come to rely on. Therefore, pre-flood self-care provisioning should be targeted

Beyond self-injection, women drew on a wider self-management repertoire when health facility care was out of reach. They negotiated traditional and calendar-based family planning with their partners and treated infection with herbal preparations and sitz baths. These practices are best understood as a rational response to an absent alternative. Some are harmless methods while others are ineffective or hazardous. Their prevalence is therefore a measure of unmet need and a signal of where accessible, acceptable services are most required (13). The appropriate response is to close the referral gap by placing a reachable provider or commodity closer, and to train CHWs and TBAs to meet minimum standards of care. Our data on CHWs and TBAs show this to be feasible.

The commodity supply chain was sustained through anticipatory management, flexible last-mile transport, and redistribution. Pre-positioning before the flood peak, forecasting from documented consumption, and reserved budget lines moved much of the response upstream of the emergency. When roads failed, teams used canoes, boats, multi-modal relays, and peer-facility transfers. These strategies mirror last-mile innovations documented elsewhere in sub-Saharan Africa. Community involvement in distribution reduced stockout duration in the Democratic Republic of Congo (20), and buffer stocks and decentralized storage for CHWs in humanitarian settings (12). They also enact a recommendation from a state-of-the-field assessment of contraceptive services in humanitarian settings. That assessment argued that emergency preparedness should be built into SRH supply chains during stable periods, rather than improvised during crises (21). What sets our case apart is that these adaptations were not introduced by an outside project or pilot. They were started by the system itself, despite being under-resourced. This makes a stronger case for funding anticipatory, decentralized supply as standard practice, since a stretched system was already doing it unprompted.

The DRF gave health facilities procurement flexibility in ordinary seasons, but it broke down when surging demand and collapsing household ability to pay coincided. This echoes findings that contraceptive security in humanitarian settings remains fragile and donor-dependent even where last-mile delivery succeeds (16). Cost-recovery therefore cannot substitute for public financing of emergency SRH supply. Mechanisms such as the DRF should be externally backstopped during the predictable flood window, when they are most likely to fail. The zonal-hub and standard operating procedure reforms that participants proposed would institutionalize this protection, but they remained unrealized.

### 4.2 Infection and menstrual health during flooding

Two needs recurred once flooding began: a rise in reproductive tract and sexually transmitted infections, and difficulty managing menstruation. These patterns are consistent with reviews of menstrual hygiene management in humanitarian crises, which describe the loss of privacy, water, and supplies and the resort to improvised materials during floods (22), and with flood-specific studies from Pakistan and Bangladesh reporting the same reuse of unclean materials and its link to infection (23,24). A review of menstrual hygiene in emergencies further reported that suspected pelvic inflammatory disease was higher among displaced women using nothing or an unwashed cloth than among those using a washed cloth (25).

Women’s own understanding of the causal pathway pointed to the same conclusion. They located the infection burden in the flooded pit latrine and the contaminated water it produced, and adapted by abandoning the latrine, washing with salted hot water, drying underwear in sunlight, and purifying drinking water. The strategies that came closest to addressing these needs were anticipatory and capability-building, notably local production of reusable pads, which removed dependence on resupply during floods. Menstrual health and infection control therefore cannot be separated from water, sanitation, and hygiene. In these settings, clinical treatment alone is repeatedly overwhelmed unless the hygiene and privacy conditions that generate infection are addressed alongside it (22,25).

### 4.3 Implications for policy and practice

Our findings reframed women from passive recipients of relief into active architects of service continuity. They planned household preparedness, provisioned and administered their own contraception, managed infection and menstruation with the means available, and organized collective solutions such as contributory funds and peer groups. Where women resorted to hazardous practices, the constraint was the absence of a reachable provider rather than women’s choices (2). Flooding in this setting is recurrent and predictable. Sustaining SRH services therefore calls for treating these adaptations as routine preparedness. It means resourcing the community and women-led structures on which continuity already depends, funding anticipatory and decentralized supply, and protecting financing when household incomes collapse. Women, therefore, should be recognized as the planners of continuity, and resourced accordingly rather than routed around.

Nigeria forecasts flooding through Nigeria Hydrological Services Agency’s Annual Flood Outlook and acts ahead of it through an anticipatory action framework that already covers Fufore and Numan, releasing pre-arranged financing for health, water, sanitation, and hygiene, food, and shelter once river levels cross a trigger (26,27). SRH commodities and services should be written into that framework as a pillar, so that pre-positioning of misoprostol, intermittent preventive treatment of malaria in pregnancy with sulfadoxine-pyrimethamine (IPTp-SP), and self-injectable contraception, advance self-care provisioning, and menstrual supplies are triggered on the same forecast. This should be financed through domestic mechanisms rather than short-term projects, using the Basic Health Care Provision Fund and its emergency gateway to pre-position stock and backstop the Drug Revolving Fund during the flood window, governed through the Ward Development Committees already required to be functional (28). The community and vendor cadres that carried continuity should be paid and trained under Nigeria’s Task-Shifting and Task-Sharing Policy, which already authorizes community health extension workers and patent medicine vendors to provide injectables and support self-injection (28,29).

### 4.4 Limitations

This study drew on multiple stakeholder perspectives, enabling triangulation across women and girls, health providers, supply chain actors, and implementers. The study was conducted in communities facing recurrent rather than one-off flooding, which allowed us to examine strategies that emerge only across seasons. However, accounts of earlier seasons were subject to recall bias, and all data were self-reported and cross-sectional, capturing perceptions at a single point rather than change over time. We sampled women who had accessed SRH services in the past year, so the experiences of those who never reached care are under-represented. In addition, some strategies attributed to TBAs, Mama-to-Mama cadres, and CHWs were reported by others rather than by those actors themselves and would merit dedicated inquiry.

## 5 Conclusions

This study documented how SRH services were sustained during recurrent flooding, largely through anticipatory, community-led adaptation. Continuity depended on the combined efforts of informal structures and the formal health system, across preparation before flooding, delivery during flooding, and slower change between seasons. CHWs, TBAs, and women’s groups were key in reaching communities that health facilities could not and in supporting women’s self-care. However, workforce shortages, financial barriers, and the collapse of cost-recovery financing left much of this continuity fragile. Program design should therefore treat preparedness as routine rather than emergency response, resourcing the community and women-led structures on which continuity depends and recognizing women as architects of that continuity.

## Data Availability

All data produced in the present study are available upon reasonable request to the corresponding author.

## 6 Conflict of Interest

The authors declare that the research was conducted in the absence of any commercial or financial relationships that could be construed as a potential conflict of interest.

## 7 Author Contributions

GWK contributed to conceptualization, research design, data analysis, and drafting of the manuscript. NK contributed to conceptualization, research design, funding acquisition, and review. IB supervised fieldwork and contributed to data analysis, project administration, and review. AJA and JU supervised fieldwork and contributed to data analysis and review. FJO contributed to research design, data analysis, project administration, and drafting of the manuscript. MAB and HSM contributed to research design, validation of findings, and review. All authors read and approved the final manuscript.

## 8 Funding

This study was supported by the generous contribution of a private donor. The donor had no role in the study design, data collection, analysis, interpretation, or the decision to submit the manuscript for publication.

## 9 Abbreviations

ANC: antenatal care
CHW: community health worker
DRF: Drug Revolving Fund
FGD: focus group discussion
IPTp-SP: intermittent preventive treatment of malaria in pregnancy with sulfadoxine-pyrimethamine
KII: key informant interview
LGA: local government area
PHC: primary health care
SRH: sexual and reproductive health
STI: sexually transmitted infection
TBA: traditional birth attendant
WDC: Ward Development Committee

## 10 Acknowledgments

The authors extend appreciation to the women, girls, health providers, supply chain actors, and program implementers who gave their time to take part in this study. We are grateful to the International Rescue Committee, Adamawa field office for facilitating the fieldwork. We also thank data collection and transcription teams for their work.

## 12 Data Availability Statement

The data collected and analyzed during the current study are available from the corresponding author on reasonable request.

## Notes

### Competing Interest Statement

The authors have declared no competing interest.

### Author Declarations

The study was approved by the International Rescue Committee Institutional Review Board (H.1.00.087). It was also approved by the Modibbo Adama University Teaching Hospital, Yola Health Research Ethics Committee (MAUTHY/SUB/S.128/404). All participants provided written informed consent. Both review boards waived parental consent for married adolescents aged 15 to 17 years, who are recognized as emancipated minors in this setting.

